# Tracing the Evolution of Research on Breaking Bad News: A Global Analysis of Publications, 2000–2025

**DOI:** 10.64898/2026.09.14.26362941

**Authors:** V Sajna Mujeeb, Lingam Ponnuchamy, Gobinda Majhi, Gyani Jail Singh Birua, Abhinith Shashidhar, Yatheesh Bharadwaj, Ancy Sara Koshy

## Abstract

**Background:** Breaking bad news (BBN) is an essential part of healthcare communication especially in the fields of oncology, palliative care and life threatening illness.

**Objective:** This study aims to analyze the global research trends, publication patterns and the intellectual structure of BBN literature from 2000 to 2025.

**Methods:** Bibliometric study was performed on Scopus database. A total of 843 publications were retrieved and analyzed by bibliometric techniques. Visualization tools such as VOSviewer and R Studio were employed, along with Lotka’s and Bradford’s laws.

**Results:** The findings indicate that there is a steady growth of publications. The United States emerged as the leading contributor. Journals including Patient Education and Counseling were dominant in the field. Key areas of focus identified through thematic analysis included oncology, palliative care and communication, while emerging themes included empathy and telemedicine.

**Conclusion:** BBN research has evolved into a mature, interdisciplinary field with increasing emphasis on patient-centered communication.

## INTRODUCTION

Breaking bad news is a critical and sensitive component of healthcare communication, particularly in clinical settings involving life-threatening illnesses, chronic conditions, and end-of-life care. It refers to the process by which healthcare professionals convey information that negatively alters a patient’s perception of their future, such as diagnosis of serious illness, poor prognosis, or treatment failure. Effective communication of bad news is not only a clinical responsibility but also an ethical obligation, as it significantly influences patients’ psychological well-being, decision-making, treatment adherence, and overall quality of care.

During the last decades, patient-centered communication has increasingly been recognized as important, resulting in the development of structured protocols (e.g., the SPIKES model, Baile et al., 2000) and other evidence-based frameworks. These approaches emphasize empathy, clarity and shared decision making in addressing the emotional and informational needs of patients and their families. But these developments are significant, but health professionals still find it hard to deliver bad news because of cultural differences, the emotional weight of the situation, lack of formal training and time pressures in clinical practice (Aljubran, 2012).

The field of breaking bad news is inherently interdisciplinary, spanning areas such as oncology, palliative care, psychiatry, medical education, and health communication. Research in this area has grown considerably as healthcare systems become more complex and there is an increased emphasis on holistic care. Different aspects of this phenomenon have been studied including communication strategies, patient preferences, ethical dilemmas, training interventions and psychosocial outcomes. But the rapid development of literature requires a systematic assessment to understand the evolution, trends and knowledge structure of the field.

Bibliometric analysis is a robust method for measuring research productivity, recognizing impactful studies, delineating collaboration networks, and uncovering emerging themes. The use of bibliometric techniques has been shown to be a useful tool to understand the development of a research domain, main contributors and patterns of scientific dissemination. Such analyzes are particularly helpful in fields like breaking bad news that span several disciplines and journals.

The current study aims to conduct a comprehensive bibliometric analysis of global research tendencies in breaking bad news using the Scopus database. This study attempts to provide a holistic picture of the intellectual landscape of the field by analyzing publication patterns, citation dynamics, authorship networks and thematic evolution over a 25-year period (2000-2025). The findings are expected to inform future research, foster interdisciplinary collaboration, and assist in the development of effective communication practices in healthcare.

## METHODS AND MATERIALS

### Aim

To explore the Global Research trends in the field of breaking Bad News.

### Objectives

1. To assess the Annual growth rate related to Research in the field of breaking the bad news
2. To assess the Institute-wise, Journal-wise, Country-wise distribution of publications.
3. To map the co-authorship networks in the field of Breaking the bad news
4. To map the Citation and co-citations networking
5. To explore the clusters associated with breaking the bad news
6. To perform the Author’s productivity and applicability of Lotka’s law.
7. To explore the zones of journals using Bradford zone and core journals related to breaking the bad news

#### Research design

The present study followed Quantitative Method. In the Initial phase, the researcher used a Descriptive approach for year-wise, country-wise, journal-wise, author-wise, and institution-wise distribution of the publications. In the second phase, an analytical approach was followed, where the researcher focused on co-authorship analysis, co-citation analysis, and Co-occurrence analysis.

#### Sources of Data Collection and data acquisition

The first step of any Bibliometric Analysis is to acquire and understand the Dataset. Scientific mapping requires a systematic approach to data acquisition. The data was acquired using the Scopus Database. The Scopus database has a wide range of journals and publications than the Web of Science and has 20% more coverage than the latter one.

#### Keyword Search Strategy

The research team conducted an extensive search to retrieve the publications. The Search Strategy was based upon the Boolean Search model. The following was the keyword search strategy used to extract the data-

TITLE (“breaking bad news” OR “delivering bad news” OR “communicating bad news” OR “bad news communication” OR “end-of-life communication” OR “serious news disclosure” OR “SPIKES protocol” OR “disclosure of diagnosis” OR “disclosure of prognosis”) AND PUBYEAR > 1999 AND PUBYEAR < 2026 AND (LIMIT-TO (LANGUAGE, “English”)).

#### Inclusion and Exclusion Criteria

Those Publications indexed in Scopus Database, published between 2000 to 2025, specifically in English Languages were included. The publication was taken for bibliometric analysis only if the title has breaking the bad news related terms.

#### Data collection process

After the inclusion and exclusion criteria and screening process, finally 843 publications were selected for the Bibliometric Analysis. The data was exported to a CSV file. The data included the following details: Authors, Title, Year, Source title, Volume, Issue, Art. No., No. of Citations, Affiliations, Author Keywords, Index Keywords, Language of Original Document, Document Type. The data was exported into CSV file on 05.03.2026.

#### Data Analysis

The data analysis for the current study was conducted using a combination of descriptive statistics, analytical techniques, and established bibliometric laws to comprehensively examine the publication trends and research patterns. Initially, Descriptive statistics were used to summarize the basic characteristics of the retrieved publications. Year-wise distribution of publications to assess temporal trends in research output, Journal-wise distribution to identify the most productive and influential journals, Institution-wise distribution to determine leading research organizations, Country-wise distribution to explore global research contributions, Types of publications (e.g., articles, reviews, conference papers) to understand the nature of scientific output. Later, to explore the intellectual structure and collaboration patterns within the field, co-authorship analysis to examine patterns of collaboration among authors, institutions, and countries, Co-citation analysis to identify influential authors, documents, and journals forming the knowledge base of the field, Keyword analysis (co-occurrence) to detect major research themes, trends, and emerging topic. To further validate the scientific productivity and distribution patterns, classical bibliometric laws such as Lotka’s law and Bradford law were used. To analysis the data and performing the visualization, bibliometric softwares such as VOSviewer and R Studio were used.

## 3. RESULT

The table 1 gives a comprehensive overview about the literature related to Breaking bad news from 2000 to 2025 using Scopus database. The time span covers 843 publications derived out of 507 journals, books and other publication types. The data demonstrates that the annual growth rate of publications is 7.43% which indicates that there is a steady increase in the research output over the period of 25 years. The average age of the documents was 9.66 years. The average number of citations per document was 21.55. The total number of references across all documents was 4,376. The dataset contained 2,349 Keywords Plus (ID) and 1,227 author-provided keywords (DE). A total of 2,874 authors contributed to the dataset. Among them, 142 authors were associated with single-authored documents. There were 148 single-authored documents. The average number of co-authors per document was 4.05. The percentage of international co-authorships was 14.35%. The dataset included multiple document types. These comprised 584 articles, 98 reviews, 47 book chapters, 42 letters, 30 notes, 17 editorials, and 13 conference papers. Additional document types included 6 short surveys, 4 errata, and 2 books.

**Table 1.** Main Information of the study.

| Description | Results |
| --- | --- |
| Timespan | 2000:2025 |
| Sources (Journals, Books, etc) | 507 |
| Documents | 843 |
| Annual Growth Rate % | 7.43 |
| Document Average Age | 9.66 |
| Average citations per doc | 21.55 |
| References | 4376 |
| **DOCUMENT CONTENTS** |  |
| Keywords Plus (ID) | 2349 |
| Author's Keywords (DE) | 1227 |
| **AUTHORS** |  |
| Authors | 2874 |
| Authors of single-authored docs | 142 |
| **AUTHORS COLLABORATION** |  |
| Single-authored docs | 148 |
| Co-Authors per Doc | 4.05 |
| International co-authorships % | 14.35 |

### 3.1 Annual scientific production of Research

The Figure 1 represents the annual scientific production which included publications from 2000 to 2025. In 2000, there were 8 articles, followed by 20 articles in 2001, 14 in 2002, 16 in 2003, and 13 in 2004. The number of articles was 15 in 2005, 16 in 2006, 17 in 2007, 15 in 2008, and 21 in 2009. In 2010, there were 24 articles, followed by 23 in 2011, 27 in 2012, 33 in 2013, and 23 in 2014. The dataset contained 35 articles for 2015, 30 for 2016, 49 for 2017, 44 for 2018, and 47 for 2019. In 2020, there were 49 articles, followed by 60 in 2021, 71 in 2022, 64 in 2023, 61 in 2024, and 48 in 2025.

**Figure 1.**
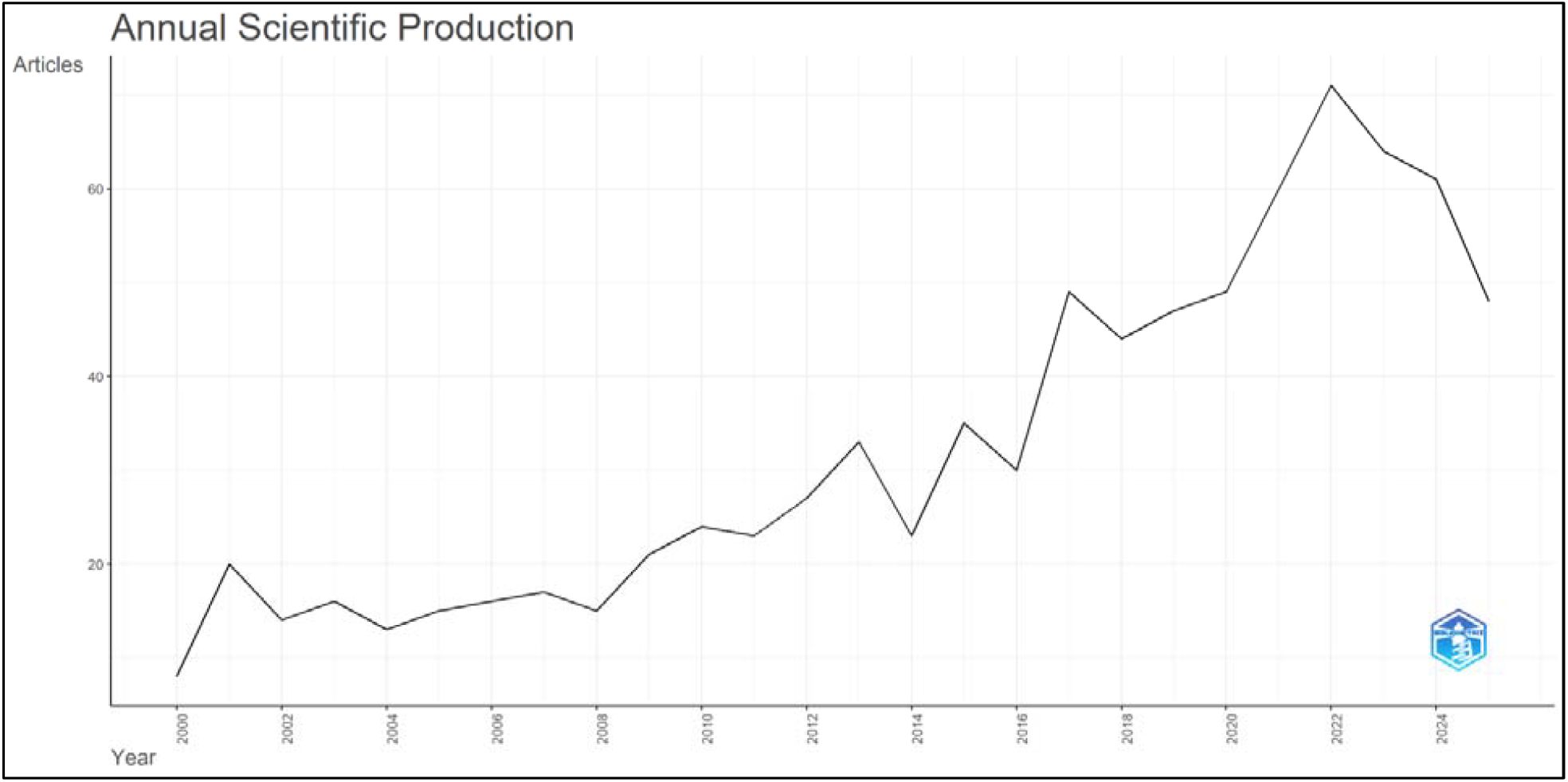
represents the annual scientific production

### 3.2 Average Annual Citations

The figure 2 represents the annual average citations. The mean total citations per article (MeanTCperArt) were highest in 2000 (311.38), followed by 2005 (71.20) and 2007 (61.24). The following years present moderate values and recent years have lower values, with 3.16 in 2024 and 0.65 in 2025, showing limited accumulation of citations. The number of articles (N) showed a steady increase over time, with a higher number of publications recorded in recent years, for example, 71 in 2022, 64 in 2023 and 61 in 2024. Mean total citations per year (MeanTCperYear) were higher in early years with 11.53 in 2000 and 3.24 in 2005. Relatively lower values were found in recent years including 0.32 in 2025.

**Figure 2.**
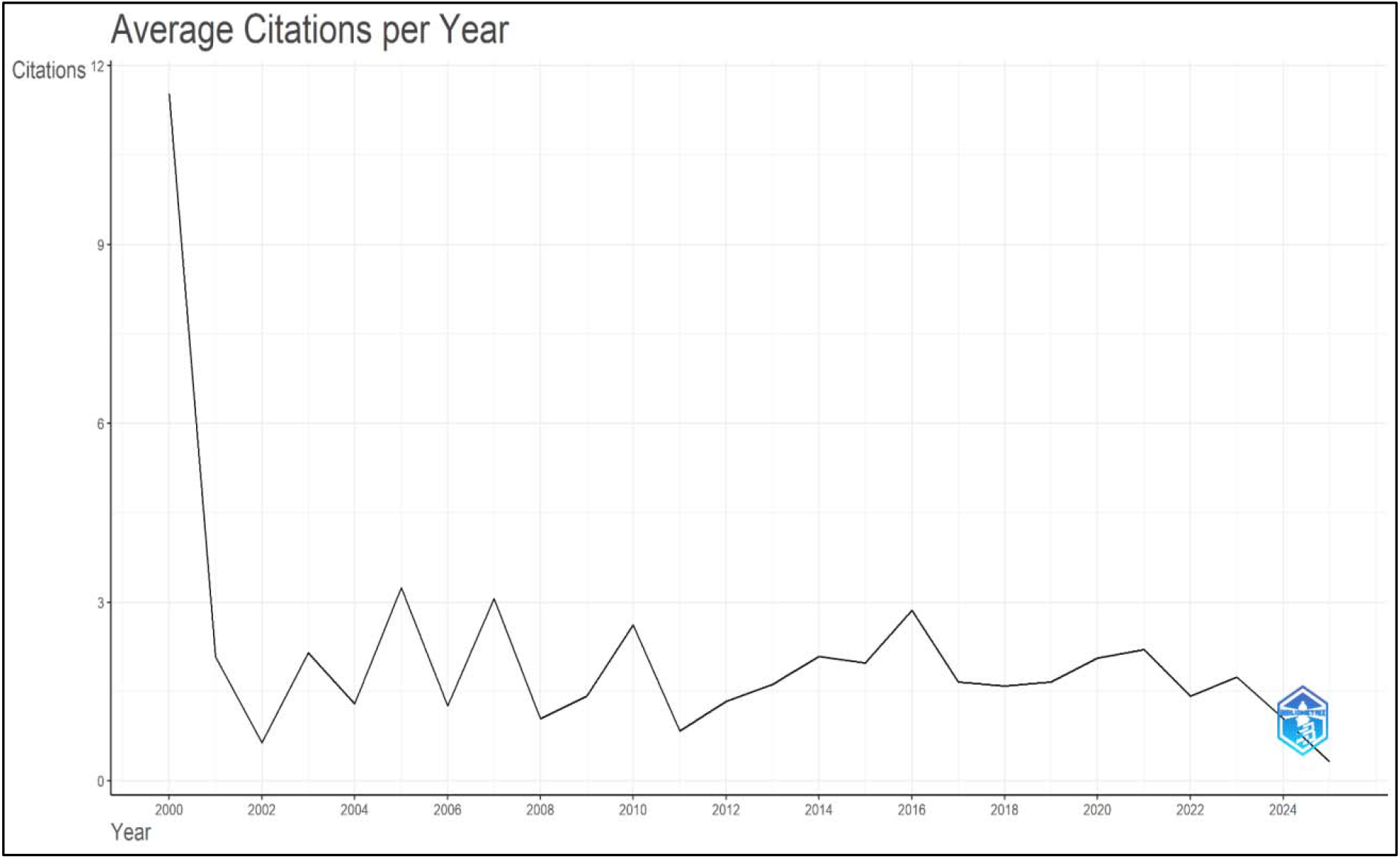
represents average annual citations

### 3.3 Projected Life Cycle of Annual Publications

The figure 3 represents the life cycle and cumulative growth curve of the annual publications. The publication trend in breaking bad news research follows a logistic growth model with strong fit (R^2^ = 0.891), indicating a systematic increase in scholarly output over time. The field grew slowly in the early years but exploded between 2015 and 2025, reflecting the increasing recognition of communication as an essential component of patient-centered care. The model predicts a peak of about 64 publications at about 2027.8, after which a decline begins, which could be a sign of a trend toward maturity. The cumulative growth curve shows an estimated saturation level of 2088 publications, with an S-shaped curve reflecting early development, rapid expansion and gradual stabilization. Such findings suggest that the field is nearing intellectual maturity and that future work will likely focus on refinement, contextual adaptation, and application-oriented studies.

**Figure 3.**
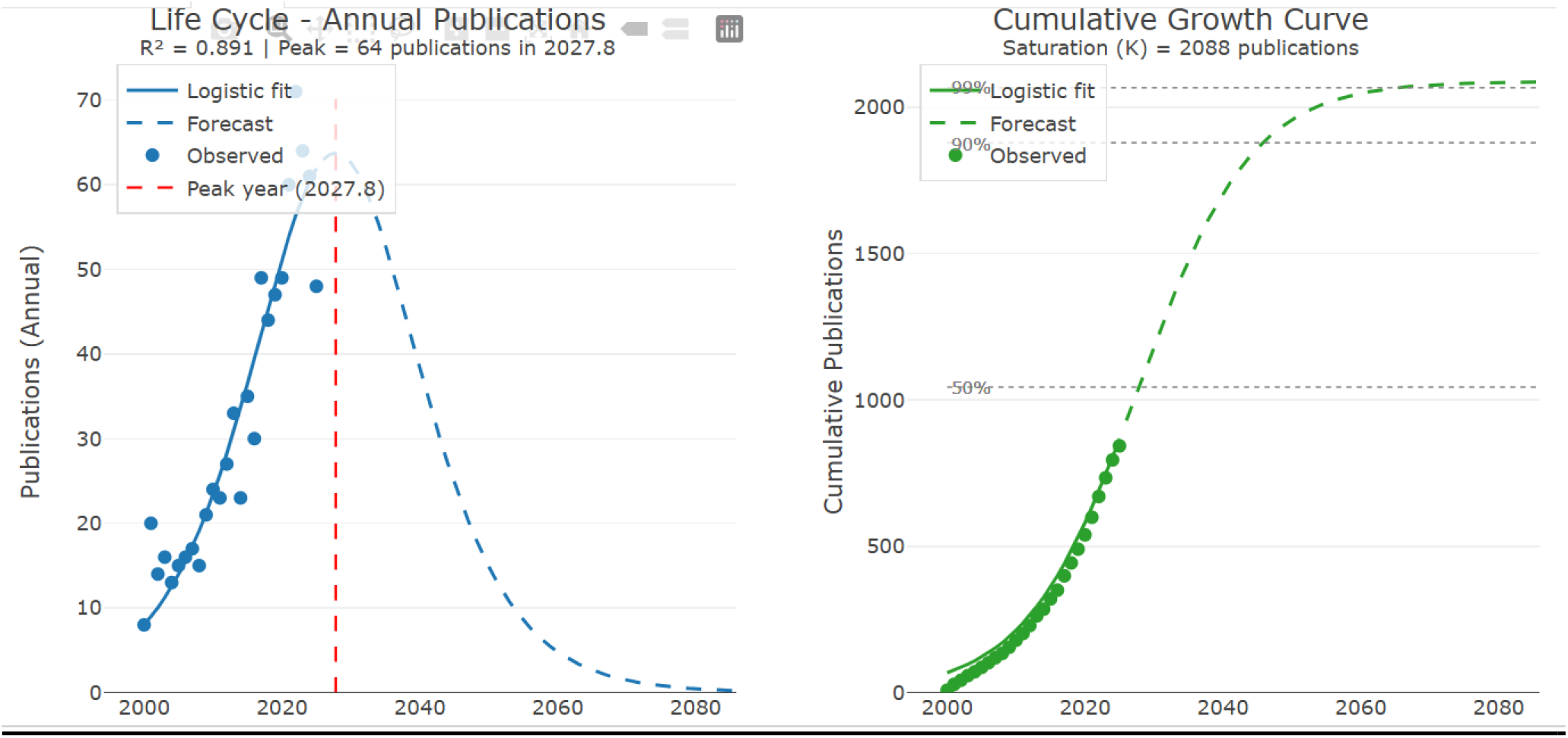
represents life cycle & cumulative growth curve of the annual publications

### 3.4 Source Analysis

The figure 4 represents the source analysis. The most relevant sources reveals that Patient Education and Counseling is the leading journal in breaking bad news research, contributing the highest number of publications (n = 40), highlighting the centrality of patient communication and counseling in this field. The Journal of Palliative Medicine (n=15) and the Journal of Cancer Education (n=13) follow, reflecting the prominence of oncology and palliative care contexts. Journals such as Health Communication and Psycho-Oncology further highlight the psychosocial and behavioral aspects of delivering bad news. The existence of so many palliative care and supportive oncology journals suggests that the field is well established in clinical practice, particularly when dealing with a life-limiting illness.

**Figure 4.**
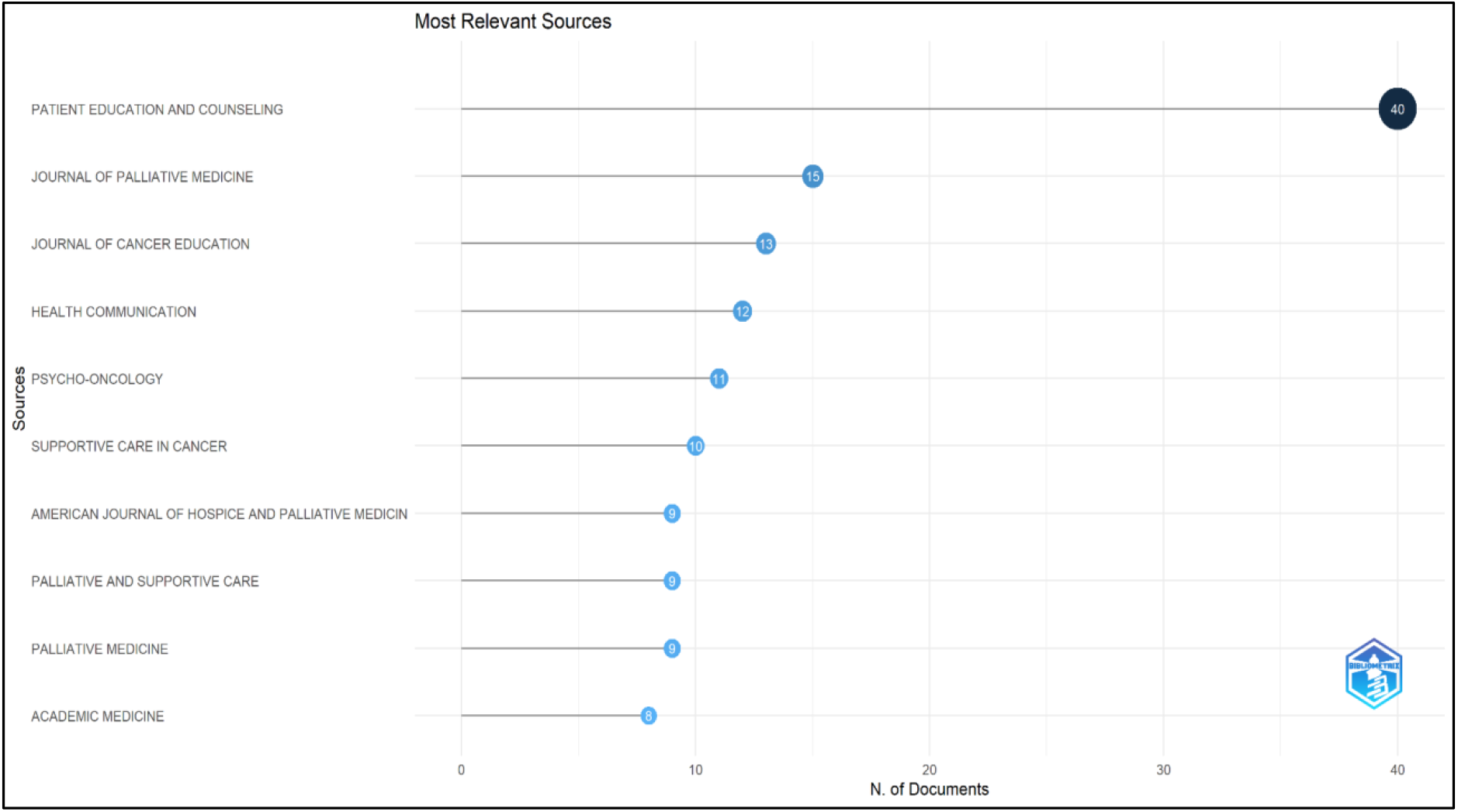
represents the source analysis

**Figure 5.**
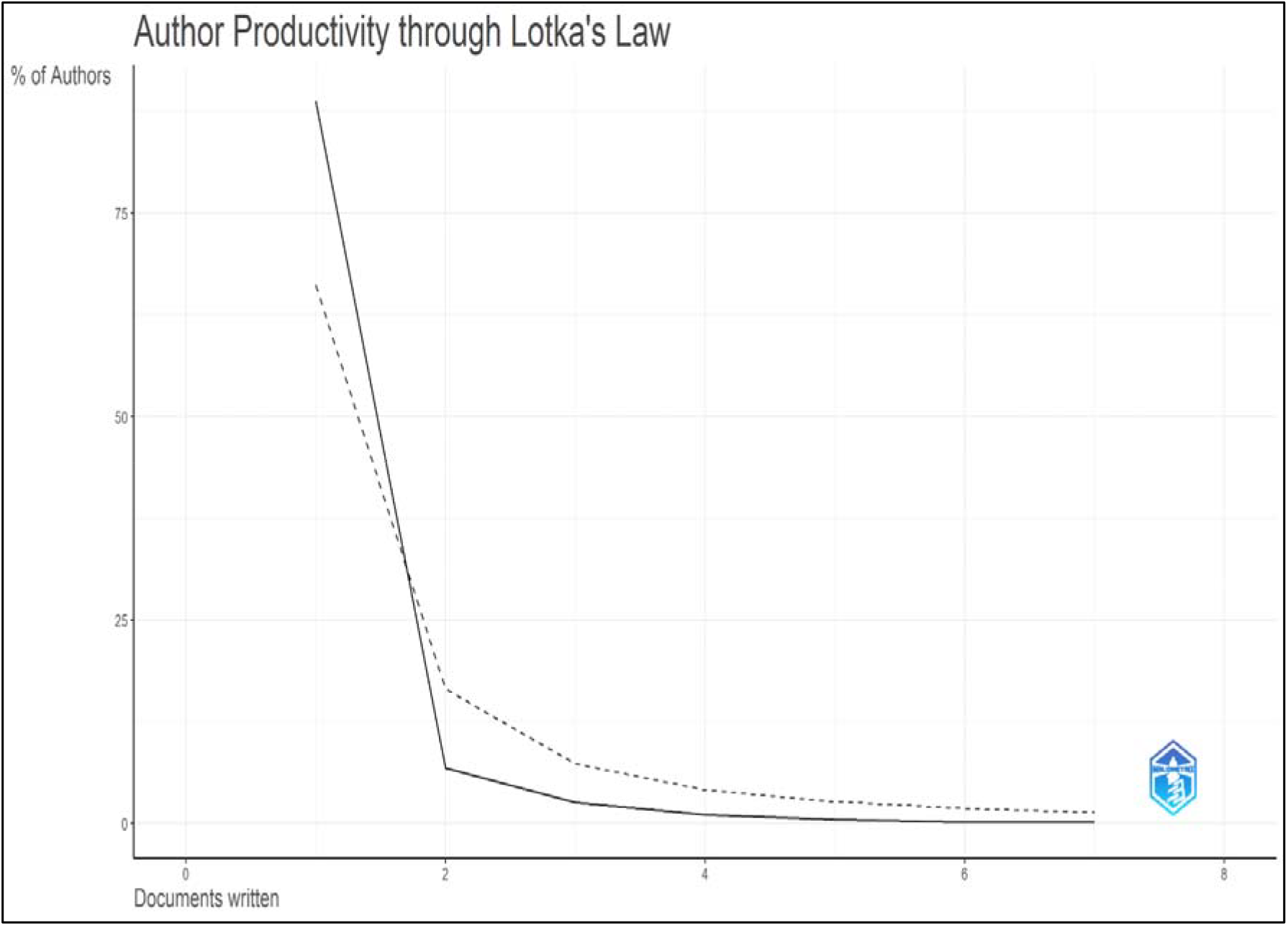
represents the Lotka’s law

**Figure 6.**
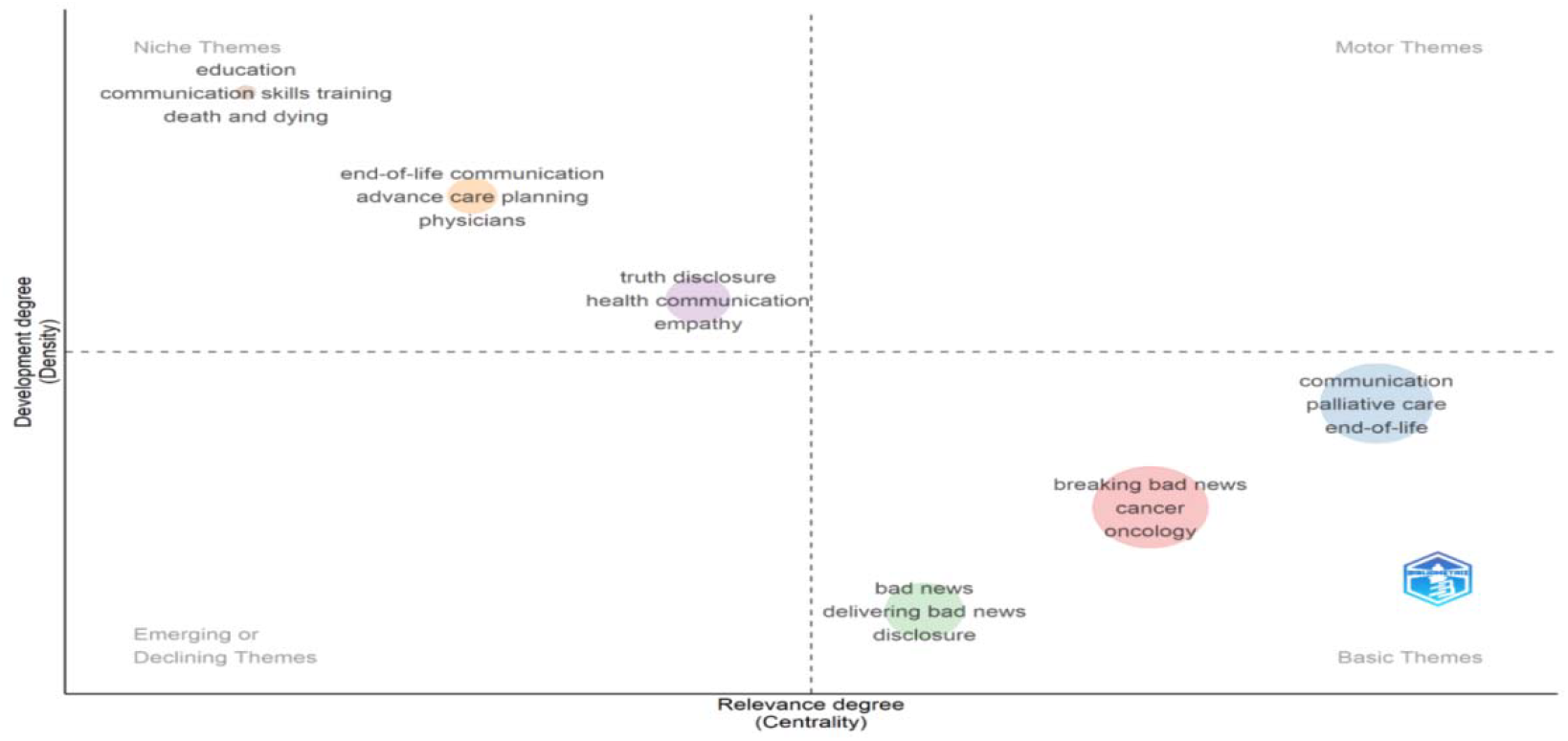
represents the thematic mapping related to breaking bad news research

### 3.5 Application of Bradford law

Bradford law is shown in Table 2 As shown in table 2, the application of Bradford’s Law indicates that a relatively small number of core journals (n = 35) contributed a substantial proportion of the total publications (33.1%), highlighting the presence of highly specialized and influential sources in breaking bad news research. The second zone had more journals (n = 194) and a similar proportion of articles (33.6%), indicating a large spread of research across a number of disciplines. Conversely, the third zone was comprised of fewer journals (n=72) with a lower contribution (8.3%) corresponding to peripheral and less consistent sources. The distribution is in accordance with Bradford’s Law, but the lower percentage in the third zone may indicate some variation in the distribution or classification of the data.

**Table 2.** represents the Bradford law.

| Bradford Zone | Number of Journals | Number of Articles | Percentage of Articles | Description |
| --- | --- | --- | --- | --- |
| Zone 1 (Core journals) | 35 | 281 | 33.1% | Highly productive core journals contributing the largest share of publications |
| Zone 2 (Moderately productive journals) | 194 | 285 | 33.6% | Journals with moderate contribution to the research field |
| Zone 3 (Least productive journals) | 72 | 70 | 8.3% | Peripheral journals contributing very few publications |

### 3.6 Application of Lotka’s law

The distribution of author productivity as shown in the table 5 reveals a pattern that partially deviates from Lotka’s Law. A substantial majority of authors (89%) contributed only a single publication, which is considerably higher than the theoretical expectation (66%). In contrast, the proportion of authors contributing multiple publications declines more sharply than predicted, indicating a limited number of highly productive contributors. This suggests a fragmented authorship structure, where most researchers engage with breaking bad news as a secondary area within their primary clinical or academic specialization. The findings highlight the interdisciplinary and practice-driven nature of the field, characterized by widespread but infrequent contributions rather than sustained author dominance.

### 3.7 Countries production of publications

The country-wise distribution of publications reveals that the United States is the leading contributor to breaking bad news research, followed by the United Kingdom, Canada, and several European countries. This dominance reflects the strong emphasis on patient-centered care, communication training, and well-established research infrastructure in high-income settings. European countries also demonstrate consistent contributions, particularly in palliative care and ethical communication practices. Conversely, recent contributions from countries such as Iran and Brazil show an increasing interest in culturally sensitive approaches to breaking bad news. The findings imply a global but inconsistent research environment with differences in healthcare systems, cultural traditions and moral perspectives, with Western countries spearheading the development of communication frameworks and non-Western countries making contributions to contextual modifications.

### 3.8 Top 10 Global cited documents

Table 4 represents the top 10 Global cited documents. The analysis of globally cited documents highlights several landmark studies that have shaped the field of breaking bad news. The most influential work is by Baile et al. (2000), which introduced the SPIKES protocol and remains the most cited article, reflecting its foundational role in clinical communication. Other highly cited studies focus on patient preferences, communication strategies in oncology, and ethical considerations in truth-telling. Contributions from palliative care and medical education further emphasize the interdisciplinary and practice-oriented nature of the field. The high citation impact and sustained relevance of these studies indicate a strong and enduring knowledge base, with core frameworks and clinical guidelines continuing to influence research and practice.

**Table 3.** represents the top 10 most productive countries.

| Country | Number of Publications |
| --- | --- |
| USA | 646 |
| UK | 179 |
| Canada | 167 |
| Germany | 105 |
| Iran | 98 |
| Italy | 90 |
| Australia | 89 |
| France | 89 |
| Netherlands | 79 |
| Brazil | 74 |

**Table 4.** represents the Top 10 Global cited documents.

| **Paper** | **DOI** | **Total Citations** | **TC Per Year** | **Normalized TC** |
| --- | --- | --- | --- | --- |
| Baile Wf, 2000, Oncologist | 10.1634/Theoncologist.5-4-302 | 2281 | 84.48 | 7.33 |
| Hagerty R, 2005, J Clin Oncol | 10.1200/Jco.2005.11.138 | 477 | 21.68 | 6.70 |
| Parker Sm, 2007, J Pain Symptom Manage | 10.1016/J.jpainsymman.2006.09.035 | 448 | 22.40 | 7.32 |
| Parker Pa, 2001, J Clin Oncol | 10.1200/Jco.2001.19.7.2049 | 347 | 13.35 | 6.43 |
| Vandekieft Gk, 2001, Am Fam Phys |  | 266 | 10.23 | 4.93 |
| Rosenbaum Me, 2004, Acad Med | 10.1097/00001888-200402000-00002 | 247 | 10.74 | 8.34 |
| Gordon Ej, 2003, Bioethics | 10.1111/1467-8519.00330 | 239 | 9.96 | 4.64 |
| Sinuff T, 2015, J Pain Symptom Manage | 10.1016/J.jpainsymman.2014.12.007 | 187 | 15.58 | 7.87 |
| Buckman R, 2005, Community Oncol | 10.1016/S1548-5315(11)70867-1 | 177 | 8.05 | 2.49 |
| Tse Cy, 2003, Palliative Med | 10.1191/0269216303pm751oa | 161 | 6.71 | 3.13 |

### 3.9 Thematic Mapping

The thematic map depicts the structure of the research on breaking bad news, indicating the differences in thematic development and thematic relevance. Basic themes such as breaking bad news, oncology, communication and palliative care are presented as basic themes, indicating their fundamental importance but relatively low level of conceptual development. However, themes such as education, communication skills training and end of life care are niche themes. These are well-developed but specialized fields that have limited integration into the wider field. Emerging topics such as empathy, truth telling, and health communication are developing research areas that may become more prominent in the future. The relative absence of strong motor themes indicates that the field is still in a developmental stage, with opportunities for greater theoretical consolidation and interdisciplinary integration.

## 4. Discussion

The current bibliometric study presents a detailed picture of the international scientific community’s studies in breaking bad news (BBN) over a quarter century from 2000 to 2025. The results reveal a constant and stable growth in the number of publications, which can be attributed to the importance of communication in health-care provision. This positive tendency agrees with modern studies that stress the significant role of communication in patients’ satisfaction, psychological adaptation, adherence to medical therapy, and overall prognosis (Ipinnimo et al., 2025; Chaure et al., 2025; De Oliveira Lima et al., 2025). The reported growth rate (7.43%) and rising volume of publications, especially since 2015, reflect the growing attention from scholars in the research field. This is in line with the shifting trends in global health care in terms of decision making, patients’ autonomy and empathic provision of care (Sandman & Munthe, 2009; Bernacki & Block, 2014; Lakin et al., 2021; Levinson et al., 2010). In addition, this field has become more interesting as many universities in the world have now included training in communication skills in their curricula (Bylund et al., 2022; Curtis et al., 2023; Berg et al., 2021). The importance of good communication skills is underscored by recent works claiming it to be an essential clinical skill, equally important as diagnostics and procedures (Chaure et al., 2025; De Oliveira Lima et al., 2025). The analysis of the life cycle shows that the growth rate follows the S-shaped curve, which implies that the discipline is entering the stage of maturity. Such a trend coincides with the principles established by bibliometrics in describing the dynamics of scientific development. As noted by Wang et al. (2021) and Fortunato et al. (2018), new fields go through phases of intensive growth before they reach an equilibrium. Source analysis The source analysis shows that core journals such as Patient Education and Counseling, Journal of Palliative Medicine, and Journal of Cancer Education are central within the field. This is because of the close relationship between BBN and oncology and palliative medicine where talking about the diagnosis and prognosis is an integral part of the medical process. The current literature also points to the importance of oncology in BBN research due to the difficulty in talking about cancer issues and the psychological distress associated with the condition (Ferrell et al., 2020; Gilligan et al., 2021; Hemming, 2017). Conversely, the existence of journals focusing on health communication in areas such as psychology and medical education illustrates how multidisciplinary the subject is. Present-day academic work is increasingly investigating topics of a psychosocial nature, ranging from patients’ preferences, emotions involved, and cultural considerations that affect communication (Yang et al., 2025; Sisk et al., 2021; Kühner et al., 2026). For instance, it has been shown that different demographics, cultural backgrounds, and contexts determine how patients prefer to be communicated about their negative diagnoses (Bylund et al., 2022).

The observed distribution in authorship follows some characteristics of Lotka’s law, in which a substantial number of authors published one paper. The findings indicate that the field features fragmented authorship, which is a common characteristic of practice-related disciplines. Clinicians tend to publish their experiences on a sporadic basis, as opposed to continuous production, a trend prevalent in research-oriented sciences (Pao, 1985). Nevertheless, the observation of prolific authors suggests the importance of opinion leaders that have influenced practice as well as scientific research.

The database under scrutiny shows moderate co-authorship levels, averaging 4.05 co-authors per document. However, the low rate of international collaboration (14.35%) is indicative of the poor intercultural dialog in the field. Because the practice is highly culture dependent, intercultural cooperation is especially important in BBN. Recent studies have reported the differences in beliefs about the truthfulness of conversations, the need for relatives’ involvement and other cultural aspects of physician-patient communication (Cain et al., 2018; Kagawa-Singer & Blackhall, 2001). For example, research from Asia and the Middle East suggests a preference for family-centred decision-making rather than Western contexts that stress individual autonomy (Aljubran, 2012; Anuk et al., 2022). The extra contributions of countries such as Iran, Brazil and India in the present study shows the growing interest in identifying BBN practices in different cultural settings.

Another significant development in recent years is the inclusion of digital health technologies in BBN communication. The COVID-19 pandemic has resulted in increased use of telemedicine, which has brought with it new challenges in delivering bad news remotely. Further, the skills of managing emotional reactions without physical presence and ensuring privacy and clarity are required in BBN through telecommunication (Wolf et al., 2020). This is a growing area of research and is likely to increase in the next few years.

The change in the intellectual structure of the field is also revealed by the thematic mapping analysis. Core themes such as oncology, palliative care and communication remain central, but are relatively underdeveloped conceptually. Conversely, niche themes, such as communication skills training and simulation-based education, are well developed, but not well integrated into the overall research framework.

Emerging themes of more holistic and humanistic approaches echo themes of empathy, compassion, patient-centered communication, and ethical disclosure. Recent literature demonstrates that empathy is not just a “nice to have” personality trait, but a quantifiable and teachable skill that has a direct effect on patient outcomes (Alansari, 2021; Back et al, 2020; Rivet et al., 2022). Furthermore, more widespread recognition of ethical issues relating to truth-telling, hope versus realism and uncertainty have been more prevalent in BBN practice (Back et al, 2020; Polivka et al., 2024). Also, there’s more of an emphasis on the emotional toll BBN takes on the healthcare providers themselves. The delivery of bad news has been shown to cause stress, burnout and emotional exhaustion and therefore the need for supportive interventions and training programs (Alansari et al., 2020; Ravaldi et al., 2021). These problems need to be solved for the sake of patients and for sustainability of providers.

## 5. Conclusion

This bibliometric study shows that research on breaking bad news (BBN) has grown steadily worldwide from 2000 to 2025, with 843 Scopus-indexed publications and an annual growth rate of 7.43%, particularly increasing after 2015. This growth reflects the growing importance of patient-centred care, empathy, communication skills, and interdisciplinary practice in areas such as oncology and palliative care. The SPIKES protocol and communication training have contributed significantly to the development of the field. However, international collaboration remains limited, and the research is fragmented across authors and journals. The absence of strong motor themes also suggests that the conceptual development of BBN research is still limited. While the United States remains a major contributor, countries such as Iran and Brazil are increasingly contributing to the field. Future research should focus on culturally appropriate communication, digital and telemedicine-based approaches, simulation training, international collaboration, and support for healthcare professionals.

## Data Availability

All data produced in the present study are available upon reasonable request to the authors

## Acknowledgement

Nil

## Declaration of Funding

The research team declare that no funding was provided to conduct this study.

## Conflict of Interest

The research team declare no conflict of Interest

## Declaration of Artificial Intelligence

The research team declare that Quillbot was used for grammar correction.

## Declaration on Data Availability

The Authors declare that the data can be availed on request to the Corresponding author.

## Notes

### Competing Interest Statement

The authors have declared no competing interest.

